# Effects of immunosuppressive therapy reduction and early post-infection graft function in kidney transplant recipients with COVID-19

**DOI:** 10.1101/2021.06.06.21258414

**Authors:** Gaetano Alfano, Francesca Damiano, Francesco Fontana, Camilla Ferri, Francesco Giaroni, Andrea Melluso, Martina Montani, Niccolò Morisi, Lorenzo Tei, Jessica Plessi, Silvia Giovanella, Giulia Ligabue, Giacomo Mori, Giovanni Guaraldi, Riccardo Magistroni, Gianni Cappelli

## Abstract

**Background:** Kidney transplant recipients with COVID-19 are at high risk of poor outcome because of comorbidities and immunosuppression. The effects of immunosuppressive therapy reduction are unclear in patients with COVID-19.

**Methods:** We conducted a retrospective study on 45 consecutive kidney transplant recipients followed at the University Hospital of Modena who tested positive for COVID-19 by RT-PCR analysis.

**Results:** The median age of patients was 56.1 (interquartile range, [IQR] 47.3-61.1) years with a predominance of male (64.4%). Kidney transplantation vintage was 10.1 (2.7-16) years, and more than half of patients (55.6%) was on triple immunosuppressive therapy. Early reduction of immunosuppression occurred in 62.8% of patients and included antimetabolite (88.8%) and calcineurin inhibitor withdrawal (22.2%).

Of the 45 patients, 88.9% became symptomatic and 40% required hospitalization. Overall mortality was 17.8%. There were no differences in outcomes between full- and reduced-dose immunosuppressive therapy at the end of follow-up. One hospitalized patient experienced irreversible graft failure. There were no differences in serum creatinine level and proteinuria in non-hospitalized patients with COVID-19. Admitted patients had better kidney function after dismission (P=0.019). Risk factors for death were age (odds ratio [OR]: 1.19; 95%CI: 1.01-1.39), and duration of kidney transplant (OR: 1.17; 95%CI: 1.01-1.35). One kidney transplant recipient experienced symptomatic COVID-19 reinfection after primary infection and anti-SARS-CoV-2 mRNA vaccine.

**Conclusions:** Despite the reduction of immunosuppression, COVID-19 affected survival of kidney transplant recipients with COVID-19. Age and duration of kidney transplant were independent predictors of death in COVID-19. Early kidney function was favorable in most survivors after COVID-19.

## Introduction

Since SARS CoV-2 was identified in December 2019, the pandemic spreads quickly all around the world with a disruptively impact on social and economic life. This virus yielded several new challenges to our healthcare systems that had to copes with an increase of morbidity and mortality among the most vulnerable populations.[1] Kidney transplant (KT) recipients is a subset of the population at high risk of severe COVID-19 relate consequences due to comorbidities, consequences of chronic kidney disease (CKD) and the burden of immunosuppressive therapy (IST), which in some subjects include also therapies administered before kidney transplantation to treat glomerulonephritis or underlying autoimmune diseases. [2]

Data collected so far reported that transplant recipients were at higher risk of morbidity and mortality compared to the general population [3, 4]. Despite the great emphasis on IST reduction to face the potentially lethal consequences of COVID-19, no confirming data supports its beneficial effect in terms of survival or clinical and laboratory manifestations. Additional uncertainty arises from recent literature when the tempered immune response is suggested to prevent COVID□19–induced systemic inflammatory syndrome leading acute respiratory distress syndrome and sepsis. Furthermore, few data are available regarding early graft outcomes after COVID-19 in survivors.[5]. Multiple causes, including kidney tropism of SARS-CoV-2 [6] may be responsible for non-reversible episodes of AKI[7] that can severely impair graft survival. Lastly, a concerning issue is hyporesponsiveness to anti-SARS-CoV-2 vaccination[8, 9]. Numerous studies have confirmed that KT recipients have a blunted immune response to mRNA vaccine[10]. Only 48% of patients was able to mount a protective serologic response to SARS-CoV-2[11]. Caillard et al [12]reported that about one-third of kidney transplant patients had severe manifestations, including a fatal outcome, after completing vaccination with mRNA. This group of patients is therefore expected to remain vulnerable to the severe complications of COVID-19 until new strategies will be implemented to reduce the susceptibility of these subjects.

Considering all the uncertainties in the management of KT recipients and the still current risk of infection in this cohort of patients we report our experience in managing KT recipients with COVID-19. In particular, we will focus on the impact of early IST reduction and early graft function after the resolution of the infection.

## Material and methods

### Kidney transplant outpatient clinic

This kidney transplant outpatient clinic follows more than 500 KT recipients, including combined liver and pancreas-kidney transplantation. Outpatient service was delivered by a senior nephrologist with experience in kidney transplantation, one fellow and three nurses. A 24-h, 7/7 days per week service was available for KT recipients in case of kidney-related pathologic process (anuria, fluid overload) or infections. This service was also offered to the subjects transplanted in our Center but living far from it.

During COVID-19 all the patients were instructed to call the clinic in case of COVID-19 symptoms. Despite the reduction of non-essential healthcare services, our outpatient clinic continued to deliver care for KT recipients adopting all the containment measures (triage at entry, masking, social distance and cleaning hands) to prevent COVID-19 diffusion. A telephonic triage was performed for all patients before reaching the hospital to intercept some paucisymptomatic patients. Patients with symptoms were invited to perform RT-PCR on nasal swab and were visited in a dedicated room to assessed vital parameters and clinical conditions. According to the severity of symptoms, patients were sent home or to the emergency room. To reduce the workload to the emergency room, patients were managed as an outpatient unless they developed severe symptoms requiring hospital admission. Remote patient monitoring was performed principally with phone calls and email.

According to our internal protocol and taking into account European expert opinions[13, 14], immunosuppressive was modulated as follow:

‐ for asymptomatic or mild COVID-19 (e.g., mild upper respiratory and/or gastrointestinal symptoms, temperature < 38°C without dyspnea) in patients on triple therapy (calcineurin-inhibitors [CNI] + mycophenolate acid [MPA]/ azathioprine [AZA] + steroids), MPA or AZA was withdrawn, and a dual therapy (CNI + steroid) was continued. If patient the patient was on dual therapy (CNI+ mammalian target of rapamycin inhibitor [MPA/mTOR-i]), MPA/mTOR was withdrawal and replaced with a low dose of steroids.
‐ for moderate and severe COVID-19 all immunosuppressors but steroid were stopped

### COVID-19 population

The study population was comprised of kidney transplant recipients with COVID-19 with a complete follow-up including death or discharge from hospital. All patients were followed in the nephrology outpatient clinic at the University Hospital of Modena. This outpatients clinic delivers care to all patients after three months from kidney transplantation.

We retrospectively reviewed the electronic charts of all KT recipients with COVID-19 from March 7, 2019, to June 25, 2021. The diagnosis of COVID-19 was performed through reverse transcriptase-polymerase chain reaction (RT-PCR) assay on a nasal swab. We excluded patients aged<18 years. Kidney function was estimated by glomerular fraction rate (eGFR) using CKD-EPI equation. Missing occurred for patients admitted to a hospital located far from our Center.

This study has been authorized by the local Ethical Committee of Emilia Romagna (n. 839/2020). The study protocol complies with the guidelines for human studies and includes evidence that the research was conducted ethically in accordance with the World Medical Association Declaration of Helsinki.

### Statistical analysis

Baseline characteristics were described using median (interquartile range [IQR]), mean and standard deviation (SD), or frequencies, as appropriate. The chi-square or Fisher’s test, and student’s t-test were used to compare categorical and continuous variables between groups, respectively. Univariate and multivariate logistic regression were performed to test the association between mortality and baseline patient characteristics. Variables that were significant on univariate analysis (P =<0.1) were entered into the multivariate model to identify independent predictors (P= < 0.05). Results were expressed as odds ratios (OR) and 95% confidence intervals (CI). Univariate and multivariate logistic regression analysis determined risk factors for death.□A P value of <0.05 was considered statistically significant. All statistical analyses were performed using SPSS® statistical software.

## Results

From the spread of COVID-in Italy, 45 KT recipients followed in at our Center contracted COVID-19. The demographic and clinical characteristics of the patients were detailed in Table 1. This group of patients included two (4.4%) combined liver-kidney and one (2.2%) heart-kidney transplant recipient.

**Table 1.**
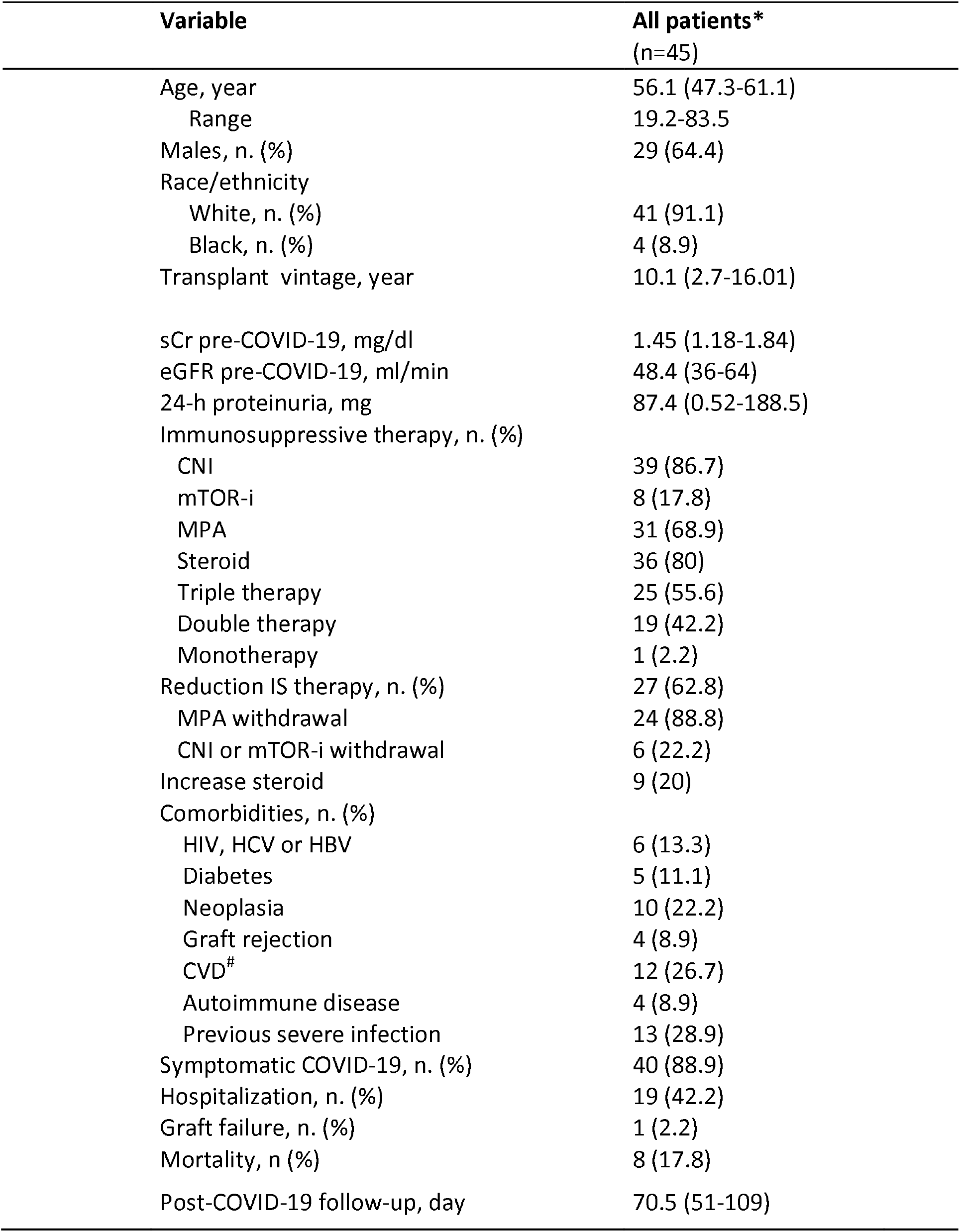
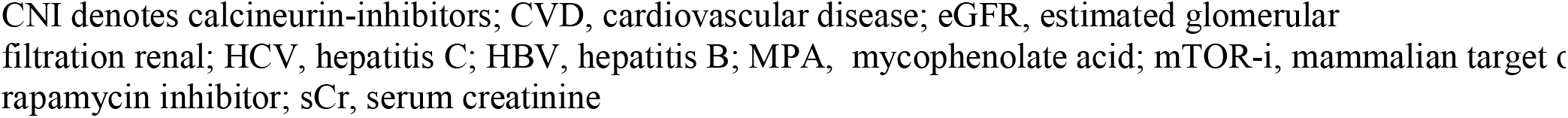
Demographics and clinical characteristics of KT recipients

Age of patients ranged from 19.2 to 83.5 years and median was 56.1 (IQR, 47.3-61.1) years. COVID-19 was prevalent in males (64.4%) and occurred after a median of 10,1 (2,7-16,01) years from transplantation.

Before COVID-19 infection serum creatine sCr measured 1.45 (IQR 1.1-1.8) mg/dl corresponding a median eGFR of 48.4 ml/min. At the time of COVID-19 diagnosis, 55.6% of the patients was in triple standard IST. Reduction of IST was practiced in more than half (62.8) of subjects. MPA (88.8%), CNI or mTOR-i (22.2%) were the most frequent discontinued agents.

Hospitalization and death rate in patients who underwent IST reduction were 51.8% and 14.8%, respectively. However, despite IST reduction, hospitalization (P=0.71) and death (P=1) rates were not different compared to the full-dose IST group. Overall, forty patients (88.9%) developed symptoms of COVID-19 and 18 of them (45%) required hospitalization. Overall, eight patients (17.8%) died for COVID-19.

Univariate and multivariate logistic regression was performed to detect predictors of mortality (Table 2). Multivariate analysis found that age (OR=1.19 [95%CI 1.01-1.39]; P=0.034) and years spent on immunosuppressive therapy (OR=1.17 [95%CI 1.01-1.35]; P=0.040) were associated with mortality in this group of patients.

**Table 2.** Univariate and multivariate predictors of AKI through logistic regression analysis

| Variable | Univariate |  |  | Multivariate |  |  |
| --- | --- | --- | --- | --- | --- | --- |
|  | OR | CI (95%) | P-value | OR | CI 3(95%) | P-value |
| Sex |  |  |  |  |  |  |
| Male | 4.40 | 0.78 24.81 | 0.09 |  |  |  |
| Age (1-yr increase) | 1.11 | 1.02 1.22 | **0.016** | 1.19 | 1.01 1.39 | **0.034** |
| KT vintage (1-yr increase) | 1.10 | 1.00 1.21 | 0.053 | 1.17 | 1.01 1.35 | **0.040** |
| Steroid-based IST | 1.93 | 0.21 18.08 | 0.56 |  |  |  |
| Reduction IST | 1.33 | 0.26 6.869 | 0.74 |  |  |  |
| Increase of steroid | 0.52 | 0.06 4.85 | 0.56 |  |  |  |
| Triple IST | 0.51 | 0.10 2.620 | 0.42 |  |  |  |
| Double IST | 1.96 | 0.38 10.026 | 0.42 |  |  |  |
| GFR | 0.99 | 0.95 1.026 | 0.57 |  |  |  |
| GFR< 45ml/min | 1.47 | 0.32 6.80 | 0.62 |  |  |  |
| GFR 45-59 ml/min | 0.68 | 0.15 3.16 | 0.62 |  |  |  |
| sCr | 1,33 | 0,26 6.87 | 0.73 |  |  |  |
| Graft rejection | 1.52 | 0.14 16.91 | 0.73 |  |  |  |
| Autoimmune disease | 0.00 | 0.00 | 0.99 |  |  |  |
| HIV/HCV/HBV | 2.58 | 0.38 17.43 | 0.33 |  |  |  |
| Previous sever infection | 0,73 | 0.13 4.19 | 0.72 |  |  |  |
| Diabetes | 1.11 | 0.11 11.49 | 0.93 |  |  |  |
| Neoplasm | 1.12 | 0.19 6.70 | 0.89 |  |  |  |
| Cardiovascular disease | 1.73 | 0.34 8.76 | 0.50 |  |  |  |

eGFR denotes estimated glomerular filtration rate; HCV, hepatitis C; HBV, hepatitis B; IST, immunosuppressive therapy; MPA, mycophenolate acid; mTOR-I, mammalian target of rapamycin inhibitor; sCr, serum creatinine.

Among survivors (82.2%), one patient with a CKD stage 4 (GFR=20 ml/min) before SARS-CoV-2 infection, developed irreversible graft failure requiring HD. One patient (2.7%) manifested de-novo proteinuria (4100 mg/die) after resolution of COVID-19 and graft biopsy revealed IgA glomerulonephritis (available data to did not allow to classify these histological findings as de-novo or recurrent IgA glomerulonephritis). Lastly, patients experienced symptomatic reinfection with COVID-19 after primary infection and anti-SARS-CoV-2 mRNA vaccine. Early post-COVID-19 follow-up of 25 (78.3%) patients showed that pre- and post-COVID variations of sCr, eGFR and 24-hour proteinuria were not statistically significant in outpatients after resolution of COVID-19. A significantly lower sCr level (P=0.019) and eGFR (P=0.028) were measured after dismission in hospitalized patients. No differences were noted in the level of proteinuria. (Table 3)

**Table 3.** Early graft function post-COVID-19 in hospitalized and non-hospitalized KT recipients

|  | Non-hospitalized patients |  |  | Hospitalized patients |  |  |
| --- | --- | --- | --- | --- | --- | --- |
|  | Pre-COVID-19 | Post-COVID-19 | P-value | Pre-COVID-19 | Post-COVID-19 | P-value |
| sCr, mg/dl | 1.31 (1.2-1.76) | 1.33 (1.08- 1.7) | 0.85 | 1.44 (1.1-1.8) | 1.32 (1-1.7) | **0.019** |
| eGFR, ml/min | 48.8 (40.5-62.1) | 56.7 (41.5-67) | 0.25 | 48. 4 (36-64) | 56.7 (41.5-67) | **0.028** |
| 24-h proteinuria | 102 (6.2-205) | 89.4 (37.2-246.4) | 0.08 | 102 (6.2-205) | 89.4 (37.2-246.4) | 0.29 |

## Discussion

Numerous reports have alerted the scientific community regarding the unfavorable outcome of COVID-19 in patients with a reduced immune response. [1, 15] The results of this study confirmed that COVID-19 poses KT recipients at high risk of severe consequences. In our cohort of KT recipients, COVID-19 carried a higher rate of symptoms, hospitalization and mortality compared to the general population[16, 17].

We found that in a cohort of 45 KT recipients of 56.1 years with COVID-19, 40% of them developed severe symptoms requiring hospitalization. Overall mortality was 17.8%, higher than the national mortality reported in the general population, which ranges between 0.1-9.2% among countries worldwide and account for about 2% globally[18].

Since IST portends poor outcomes in patients with infections, a new therapeutic strategy was set to reduce the burden of IST in COVID-patients. In the attempt to reconstitute the immune system against SAR-CoV-2 infection, we systematically minimized anti-rejection therapy. All KT recipients, who communicated their COVID-19 positivity, were advised to withdraw at least antimetabolite agents (MFA or AZA). In hospitalized patients, IST was reduced or suspended according to the clinical conditions of the patient. Nevertheless, hospitalization and death rate in the reduced-dose IST group was not dissimilar from full-dose IST group. A potential explanation for this result is that reduction of immunosuppressive therapy was not able to reconstitute an efficient immune response after the long-term burden of IST.

Although IST reduction did not lead to a favorable outcome, it is worth mentioning that the overall mortality estimated in our cohort was tendentially lower than that reported in several monocentric and multicentric studies where this approaching up to 32.5%[19–26]. Our results are in line with the population-based data on 1013 KT recipients affected by COVID-19 collected by French and Spain national registries, which reported a 28-day mortality of 20%[27]. In Brescia (Italy), Bossini et al.[24] reported a higher overall mortality rate among KT recipients (28%) during the first wave of the infection. Similar to our therapeutic strategy, they practiced immunosuppression cessation in all hospitalized patients and introduced or increased the glucocorticoid dose. At first glance, the causes of this different mortality rate are unknown. The different timing of enrollment made the two cohorts not perfectly comparable. The enrollment of all patients of the Brescia cohort during the first wave has probably magnified the risk of side effects of therapeutic regimens administered during the first wave[28, 29] and the challenges to delivering the standard of care in an overwhelmed and unprepared hospital. Lastly, a lower age (56.1 vs 60 years) in our cohort of patients has probably contributed to lead to a better prognosis.

Multivariate analysis showed that the predictors of death were age and time elapsed on IST, in line with previous studies.

Age is widely associate with COVID-19 severity and death in KT recipients[30, 31] as well as the general population [32]. Centers for Disease Control (CDC) claims that 8 out 10 COVID-19 death in the U.S. has been reported in adults aged more 65 years and the risk of hospitalization and death increases enormously with age.[33]

The effect of immunosuppression is still controversial in KT recipients[34]. Immunosuppression is known to dysregulate innate and adaptive immunity, exposing the patients to severe infections. On the other hand, severe COVID-19 infection has been associated with a dysregulated inflammatory response (IL-6, IL-1, and chemokines) leading to ARDS and sepsis. The new insights support a promising role of immunosuppressants in the most severe case on COVID-19 (i.e., tocilizumab, steroid)[35]. In our opinion, the interplay between immunosuppression and pathogenesis of COVID-19 should be contextualized with the stage of SARS-CoV-2 infection. The high rate of symptomatic disease is probably secondary to the host susceptibility due to immunodepression and the burden of CKD. If the IST is somewhat beneficial to reduce the pro-inflammatory release of cytokines in the second phase of infection, more studies are required to elucidate which molecular pathway is involved.

Lastly, we report a short-term good graft function in survival after COVID-19. These data indicate a stable early graft function (sCr and 24-hour proteinuria) in outpatients COVID-19 who were not hospitalized. Conversely, hospitalized KT recipients had a statistically significant improvement in renal function. As supported by Dacina et al[5] we speculate that lower sCr after the COVID-19 episode was due to the minimization or withdrawn of CNI, a ‘drug holiday’ without apparently dire consequences in terms of graft rejection for 95% of survivors.

Some limitations of the study should be enunciated. It is a retrospective study, with small sample size and a short follow-up after COVID-19. The small number of patients in full- and reduced-dose of IST group may have reduced the probability to observe an underlying difference between these two groups. Furthermore, we cannot exclude that in some cases, reduction of the IST occurred after a short delay from the diagnosis of COVID-19. However, all patients with symptoms or at risk of contagious underwent nasal swab as fast as possible in an ambulatory setting.

In conclusion, reduction of immunosuppression did not reduce the risk of severe COVID-19 or death. Age and time spend on kidney transplantation are independent predictors of death in our patients. Short-term follow-up after COVID-19 showed an excellent graft function in most survivors. Primary infection or vaccination does not exclude the risk of SARS-CoV-2infection in KT recipients.

## Data Availability

Data are available upon reasonable request to the corresponding author

## Acknowledgments

Special thanks are due to Dr. Roberto Pulizzi, Francesca Facchini and Marco Ballestri skilled and experienced nephrologists involved in the “Kidney Transplant Program” and Laura Bonaretti and all nurses of the “Kidney Transplantation Outpatient Clinic” at the University Hospital of Modena for their precious support in managing KT recipients.

## Bibliography

1. Williamson EJ, Walker AJ, Bhaskaran K, et al (2020) Factors associated with COVID-19-related death using OpenSAFELY. Nature 584:430–436. https://doi.org/10.1038/s41586-020-2521-4

2. Khairallah P, Aggarwal N, Awan AA, et al (2021) The impact of COVID-19 on kidney transplantation and the kidney transplant recipient - One year into the pandemic. Transpl Int Off J Eur Soc Organ Transplant 34:612–621. https://doi.org/10.1111/tri.13840

3. Caillard S, Chavarot N, Francois H, et al (2021) Is COVID-19 infection more severe in kidney transplant recipients? Am J Transplant 21:1295–1303. https://doi.org/10.1111/ajt.16424

4. Fisher AM, Schlauch D, Mulloy M, et al (2021) Outcomes of COVID-19 in hospitalized solid organ transplant recipients compared to a matched cohort of non-transplant patients at a national healthcare system in the United States. Clin Transplant 35:. https://doi.org/10.1111/ctr.14216

5. Elec AD, Oltean M, Goldis P, et al (2021) COVID-19 after kidney transplantation: Early outcomes and renal function following antiviral treatment. Int J Infect Dis 104:426–432. https://doi.org/10.1016/j.ijid.2021.01.023

6. Cravedi P, Mothi SS, Azzi Y, et al (2020) COVID-19 and kidney transplantation: Results from the TANGO International Transplant Consortium. Am J Transplant Off J Am Soc Transplant Am Soc Transpl Surg 20:3140–3148. https://doi.org/10.1111/ajt.16185

7. Bajpai D, Deb S, Bose S, et al (2021) Recovery of kidney function after AKI due to COVID-19 in kidney transplant recipients. Transpl Int Off J Eur Soc Organ Transplant. https://doi.org/10.1111/tri.13886

8. Benotmane I, Gautier-Vargas G, Cognard N, et al (2021) Weak anti–SARS-CoV-2 antibody response after the first injection of an mRNA COVID-19 vaccine in kidney transplant recipients. Kidney Int 99:1487–1489. https://doi.org/10.1016/j.kint.2021.03.014

9. Alfano G, Fontana F, Mori G, et al (2021) Seroconversion after COVID-19 vaccine in a dialysis patient on immunosuppressants. Clin Kidney J. https://doi.org/10.1093/ckj/sfab065

10. Boyarsky BJ, Werbel WA, Avery RK, et al (2021) Antibody Response to 2-Dose SARS-CoV-2 mRNA Vaccine Series in Solid Organ Transplant Recipients. JAMA 325:2204. https://doi.org/10.1001/jama.2021.7489

11. Benotmane I, Gautier-Vargas G, Cognard N, et al (2021) Low immunization rates among kidney transplant recipients who received 2 doses of the mRNA-1273 SARS-CoV-2 vaccine. Kidney Int 99:1498–1500. https://doi.org/10.1016/j.kint.2021.04.005

12. Caillard S, Chavarot N, Bertrand D, et al (2021) Occurrence of severe COVID-19 in vaccinated transplant patients.Kidney Int 0 https://doi.org/10.1016/j.kint.2021.05.011

13. Expert-opinion-on-ISD-in-Covid-19.pdf

14. Tsalouchos A, Salvadori M (2020) La pandemia del nuovo coronavirus 2019 ed il trapianto renale. G Clin Nefrol E Dialisi 32:60–63. https://doi.org/10.33393/gcnd.2020.2133

15. Myint PK, Carter B, Barlow-Pay F, et al (2021) Routine use of immunosuppressants is associated with mortality in hospitalised patients with COVID-19. Ther Adv Drug Saf 12:2042098620985690. https://doi.org/10.1177/2042098620985690

16. Cascella M, Rajnik M, Aleem A, et al (2021) Features, Evaluation, and Treatment of Coronavirus (COVID-19). In: StatPearls. StatPearls Publishing, Treasure Island (FL)

17. Mappe Coronavirus. https://mappe.protezionecivile.gov.it/it/mappe-emergenze/mappe-coronavirus. Accessed 5 Jun 2021

18. COVID-19 Map. In: Johns Hopkins Coronavirus Resour. Cent. https://coronavirus.jhu.edu/map.html. Accessed 27 May 2021

19. Pérez-Sáez MJ, Blasco M, Redondo-Pachón D, et al (2020) Use of tocilizumab in kidney transplant recipients with COVID-19. Am J Transplant Off J Am Soc Transplant Am Soc Transpl Surg 20:3182–3190. https://doi.org/10.1111/ajt.16192

20. Aziz H, Lashkari N, Yoon YC, et al (2020) Effects of Coronavirus Disease 2019 on Solid Organ Transplantation. Transplant Proc 52:2642–2653. https://doi.org/10.1016/j.transproceed.2020.09.006

21. Coll E, Fernández-Ruiz M, Sánchez-Álvarez JE, et al (2021) COVID-19 in transplant recipients: The Spanish experience. Am J Transplant Off J Am Soc Transplant Am Soc Transpl Surg 21:1825–1837. https://doi.org/10.1111/ajt.16369

22. Akalin E, Azzi Y, Bartash R, et al (2020) Covid-19 and Kidney Transplantation. N Engl J Med. https://doi.org/10.1056/NEJMc2011117

23. Fernández-Ruiz M, Andrés A, Loinaz C, et al (2020) COVID-19 in solid organ transplant recipients: A single-center case series from Spain. Am J Transplant Off J Am Soc Transplant Am Soc Transpl Surg 20:1849–1858. https://doi.org/10.1111/ajt.15929

24. Bossini N, Alberici F, Delbarba E, et al (2020) Kidney transplant patients with SARS□CoV□2 infection: the brescia renal COVID task force experience. Am J Transplant. https://doi.org/10.1111/ajt.16176

25. Cravedi P, Mothi SS, Azzi Y, et al (2020) COVID-19 and kidney transplantation: Results from the TANGO International Transplant Consortium. Am J Transplant Off J Am Soc Transplant Am Soc Transpl Surg 20:3140–3148. https://doi.org/10.1111/ajt.16185

26. Nair V, Jandovitz N, Hirsch JS, et al (2020) COVID□19 in kidney transplant recipients. Am J Transplant. https://doi.org/10.1111/ajt.15967

27. Jager KJ, Kramer A, Chesnaye NC, et al (2020) Results from the ERA-EDTA Registry indicate a high mortality due to COVID-19 in dialysis patients and kidney transplant recipients across Europe. Kidney Int 98:1540–1548. https://doi.org/10.1016/j.kint.2020.09.006

28. Gérard A, Romani S, Fresse A, et al (2020) “Off-label” use of hydroxychloroquine, azithromycin, lopinavir-ritonavir and chloroquine in COVID-19: A survey of cardiac adverse drug reactions by the French Network of Pharmacovigilance Centers. Therapies 75:371–379. https://doi.org/10.1016/j.therap.2020.05.002

29. Izcovich A, Siemieniuk RA, Bartoszko JJ, et al (2020) Adverse effects of remdesivir, hydroxychloroquine, and lopinavir/ritonavir when used for COVID-19: systematic review and meta-analysis of randomized trials. Infectious Diseases (except HIV/AIDS)

30. Coll E, Fernández-Ruiz M, Sánchez-Álvarez JE, et al (2021) COVID-19 in transplant recipients: The Spanish experience. Am J Transplant Off J Am Soc Transplant Am Soc Transpl Surg 21:1825–1837. https://doi.org/10.1111/ajt.16369

31. Oto OA, Ozturk S, Turgutalp K, et al (2021) Predicting the outcome of COVID-19 infection in kidney transplant recipients. BMC Nephrol 22:100. https://doi.org/10.1186/s12882-021-02299-w

32. Levin AT, Hanage WP, Owusu-Boaitey N, et al (2020) Assessing the age specificity of infection fatality rates for COVID-19: systematic review, meta-analysis, and public policy implications. Eur J Epidemiol 35:1123–1138. https://doi.org/10.1007/s10654-020-00698-1

33. CDC (2020) COVID-19 and Your Health. In: Cent. Dis. Control Prev. https://www.cdc.gov/coronavirus/2019-ncov/need-extra-precautions/older-adults.html. Accessed 27 May 2021

34. Chavarot N, Gueguen J, Bonnet G, et al (2021) COVID□19 severity in kidney transplant recipients is similar to nontransplant patients with similar comorbidities. Am J Transplant 21:1285–1294. https://doi.org/10.1111/ajt.16416

35. Pérez-Sáez MJ, Blasco M, Redondo-Pachón D, et al (2020) Use of tocilizumab in kidney transplant recipients with COVID-19. Am J Transplant Off J Am Soc Transplant Am Soc Transpl Surg 20:3182–3190. https://doi.org/10.1111/ajt.16192

